# The impact of believing you have had COVID-19 on behaviour: Cross-sectional survey

**DOI:** 10.1101/2020.04.30.20086223

**Authors:** Louise E. Smith, Abigail L. Mottershaw, Mark Egan, Jo Waller, Theresa M. Marteau, G James Rubin

## Abstract

**Objectives:** To investigate whether people who think they have had COVID-19 are less likely to engage in social distancing measures compared with those who think they have not had COVID-19.

**Design:** On-line cross-sectional survey.

**Setting:** Data were collected between 20^th^ and 22^nd^ April.

**Participants:** 6149 participants living in the UK aged 18 years or over.

**Main outcome measures:** Perceived immunity to COVID-19, self-reported adherence to social distancing measures (going out for essential shopping, nonessential shopping, and meeting up with friends/family; total out-of-home activity), worry about COVID-19 and perceived risk of COVID-19 to oneself and people in the UK. Knowledge that cough and high temperature / fever are the main symptoms of COVID-19.

**Results:** In this sample, 1493 people (24.3%) thought they had had COVID-19. Only 245 (4.0%) reported receiving a test result saying they had COVID-19. Reported test results were often incongruent with participants’ belief that they had had COVID-19. People who believed that they had had COVID-19 were: more likely to agree that they had some immunity to COVID-19; less likely to report adhering to social distancing measures; less worried about COVID-19; and less likely to know that cough and high temperature / fever are two of the most common symptoms of COVID-19.

**Conclusions:** The number of people in the UK who think they have already had COVID-19 is about twice the rate of current prevalence estimates. People who think that they have had COVID-19 may contribute to transmission of the virus through non-adherence to social distancing measures. Clear communications to this growing group are needed to explain why protective measures continue to be important and to encourage sustained adherence.

**COPYRIGHT:** The Corresponding Author has the right to grant on behalf of all authors and does grant on behalf of all authors, an exclusive licence (or non exclusive for government employees) on a worldwide basis to the BMJ Publishing Group Ltd to permit this article (if accepted) to be published in BMJ editions and any other BMJPGL products and sublicences such use and exploit all subsidiary rights, as set out in our licence.

**FUNDING SOURCES:** JW is funded by a career development fellowship from Cancer Research UK (ref C7492/A17219). LS and GJR are supported by the National Institute for Health Research Health Protection Research Unit (NIHR HPRU) in Emergency Preparedness and Response at King’s College London in partnership with Public Health England (PHE), in collaboration with the University of East Anglia and Newcastle University. The views expressed are those of the authors and not necessarily those of the NHS, the NIHR or the Department of Health and Social Care, Public Health England. Data collection was funded via a block Government grant to the Behavioural Insights Team.

**COMPETING INTEREST STATEMENT:** All authors have completed the Unified Competing Interest form (available on request from the corresponding author) and declare: ALM and ME report grants from government partners to the Behavioural Insights Team, during the conduct of the study, JW reports grants from Cancer Research UK, during the conduct of the study; no financial relationships with any organisations that might have an interest in the submitted work in the previous three years, no other relationships or activities that could appear to have influenced the submitted work.

**TRANSPARENCY DECLARATION:** The authors affirm that the manuscript is an honest, accurate, and transparent account of the study being reported; that no important aspects of the study have been omitted; and that any discrepancies from the study as originally planned have been explained.

**AUTHOR CONTRIBUTION STATEMENT:** The study was conceptualised by LS, GJR, JW and TMM. AM and ME completed data collection. LS analysed the data. All authors contributed to, and approved, the final manuscript.

**WHAT IS ALREADY KNOWN ON THIS TOPIC:**

- During the COVID-19 pandemic, multiple countries, including the UK, have introduced “lockdown” measures.
- The World Health Organization has warned against using the results of antibody tests to issue “immunity passports” due to fears that those who test positive for antibodies may stop adhering to protective measures.
- There is no research investigating adherence to protective measures among those who think they have had COVID-19.

**WHAT THIS STUDY ADDS:**

- This is the first study investigating behavioural differences between those who think they have had COVID-19 and those who do not.
- About twice as many people think they have had COVID-19 than prevalence estimates suggest.
- Results suggest that there may be a high degree of self-misdiagnosis within those who think they have had COVID-19.
- Those who think they have had COVID-19 were more likely to think they were immune to COVID-19, and less likely to adhere to social distancing measures.

## INTRODUCTION

Since the onset of the COVID-19 outbreak, numerous countries have introduced “lockdown” measures to limit contact between people and reduce the spread of illness. Some reports estimate that 3.9 billion people, over half the world’s population, have been asked to “stay at home” by their country’s Government or have had some limit on their movement imposed.(1) With almost 3 million confirmed cases globally,(2) countries around the world are pinning their hopes on testing as part of their exit strategy from more severe measures. This includes antigen testing, which tests if people currently have COVID-19, and antibody testing, which tests if people have had COVID-19 in the past. However, the World Health Organization has warned against using antibody tests in order to issue people with “immunity passports” due to fears that those who test positive for antibodies may stop adhering to protective measures.(3)

People who believe they have had COVID-19 may be more likely to think they are completely immune, stop engaging in protective behaviours such as handwashing and reduce their social distancing measures. This may contribute to transmission of the virus for two reasons. First, test results can be wrong (4) and in the absence of testing people can misdiagnose themselves: this can lead people to believe that they have had COVID-19 when they have not. Second, for people who have had COVID-19, it remains unknown whether they could catch COVID-19, and be infectious, more than once.(3) However, there is currently no evidence about the behavioural consequences of believing that you have had COVID-19.

In this study, we explored whether believing that you have already had COVID-19 alters your behaviour. We hypothesised that people who think they have had COVID-19 are: more likely to believe that they are immune to COVID-19; less likely to adhere to social distancing measures; less worried about COVID-19; and perceive a lower risk of COVID-19 to themselves, but no difference in perceived risk of COVID-19 to others. We also investigated awareness of the most common COVID-19 symptoms as a marker of likely accuracy of self-diagnosis.

## METHOD

### Design

This cross-sectional survey was carried out by the Behavioural Insights Team on their in-house online experimentation platform, Predictiv, between 20^th^ and 22^nd^ April.

### Participants

Participants (n=6149) were recruited from Predictiv’s research panel (n=500,000 UK adults) and were eligible for the study if they were aged 18 or over and lived in the UK. Quotas were based on age, gender, income and region to ensure a sample that was broadly representative of the general UK population. 89% of people who clicked on the link subsequently completed the study materials. For this survey, participants were reimbursed in points (equivalent to up to approximately £1) which could be redeemed in cash, gift vouchers or charitable donations. Participants did not know the topic of the survey before commencing it.

### Study materials

These questions were asked as part of an experimental study investigating behavioural outcomes of antibody test terminology.(5) Results of this experiment will be reported elsewhere. For the purposes of this paper, we collapsed the data across all arms of the experiment and controlled for experimental condition.

Having had COVID-19 Participants were asked “Do you think you have already had coronavirus?” Response options were “Yes, definitely”, “Yes, probably”, “No, probably not”, and “No, definitely not”.

### Other measures

We asked participants if they had been tested for COVID-19. Possible answers included “yes, the results showed I did have coronavirus”, “yes, the results showed I did not have coronavirus” and “no, I haven’t been tested”.

To measure perceived immunity to COVID-19, we asked participants to what extent they agreed or disagreed with the statement “I think I have some immunity to coronavirus” on a five-point Likert scale (“strongly agree” to “strongly disagree”).

We asked participants to state “over the last seven days, on how many days” they had: been to the shops, for groceries/pharmacy, been to the shops, for things other than groceries/pharmacy, gone for a walk or some other exercise; gone out to work, helped or provided care for a vulnerable person, and met up with friends and/or family they did not live with.

We asked participants to rate how worried they were about COVID-19 on a five-point Likert-type scale from “not at all worried” to “extremely worried”. We also asked participants to rate the extent to which they thought COVID-19 posed a risk to themselves personally and to people in the UK on a four-point Likert-type scale from “no risk at all” to “major risk”.

To assess the likelihood of misdiagnosis, we asked participants what they thought “the most common symptoms of coronavirus” were from a list of thirteen items (including cough, high temperature / fever, shortness of breath / difficulty breathing, runny or blocked nose, aches and pains, chest pain, chills / shivering, sore throat, diarrhoea, headaches, stomach ache, feeling tired or having low energy, and loss of sense of smell / taste). Participants could select up to three symptoms.

### Personal characteristics

Participants were asked to state their: age; gender; employment status; highest educational attainment; and region. Participants were also asked what sector they worked in (to identify key workers) and whether they had children.

### Ethics

Ethical approval for this study was granted by the King’s College London Research Ethics Committee (reference: MRA-19/20-18485).

### Patient and public involvement

Due to the rapid nature of this research, the public was not involved in the development of the survey materials.

### Power

A sample size of 6,150 allows a 95% confidence interval of plus or minus 1.25% for the prevalence estimate for each survey item.

### Analysis

We recoded thinking you have had COVID-19 into a binary variable (yes / no), grouping together responses of “Yes, definitely” and “Yes, probably”, versus “No, probably not” and “No, definitely not”.

We created a binary variable to identify whether participants had correctly identified cough and high temperature / fever as two out of the three most common symptoms of COVID-19. We coded those who answered “don’t know” as incorrect.

We defined non-adherence to social distancing measures by considering the instructions from the UK Government to members of the public that were in force at the time of data collection.(6) If participants went out to the shops for items other than groceries/pharmacy once or more in the last seven days, or met up with friends and/or family they did not live with once or more in the last seven days, we classed them as not adhering to the guidelines. There is no objective guidance on the frequency of shopping for basic necessities such as food or medicine, with guidance stating that it “must be as infrequent as possible”.(7) We created a binary variable for shopping for groceries/pharmacy grouping together those who had been shopping for necessities on two or more days in the last week, compared to one day or less. We also created a continuous variable representing the total amount of out-of-home activity a participant had engaged in during the past week, by summing the number of days they had left the house for each of six activities (shopping for groceries/pharmacy, shopping for items other than groceries/pharmacy, going for a walk or some other exercise, going out to work, helping or providing care for a vulnerable person; meeting up with friends and/or family they did not live with).

For all analyses with binary outcomes (correct identification of the most common symptoms of COVID-19; non-adherence to social distancing measures), we used binary logistic regressions to investigate univariable associations between thinking you have had COVID-19 and dependent variables. We then used a second logistic regression adjusting for all personal characteristics and experimental group.

For analyses with a continuous outcome (perceived immunity to COVID-19; worry about COVID-19; perceived risk of COVID-19 to oneself; perceived risk of COVID-19 to people in the UK; out-of-home activity), we used a series of one-way ANOVAs to investigate univariable associations between thinking you have had COVID-19 and dependent variables. We then used a series of ANCOVAs adjusting for all personal characteristics and experimental group.

Our analyses report unweighted statistics. We corrected for multiple comparisons using a Bonferroni adjustment (*p*=.005).

To provide a graphical illustration of the results, we used a bar chart to show the differences between those who did and did not think they had had COVID-19 in terms of the proportions giving responses at the extreme end of the scale for relevant outcomes (e.g. strongly agreeing they have some immunity, being not at all worried about COVID-19).

### Sensitivity analyses

We re-ran analyses excluding those who had been tested for COVID-19.

## RESULTS

Only adjusted analyses are reported narratively; unadjusted analyses are reported in the tables.

### Participants

24.3% (n=1493) of participants thought that they had had COVID-19. 9.4% (n=575) participants had been tested for COVID-19. Of those who had been tested, 42.6% (n=245) reported that the test showed they did have COVID-19, while 57.4% (n=330) reported that the test showed they did not have COVID-19. Of those who reported that their test showed they did not have COVID-19, 56.7% (n=187) nonetheless thought that they had had COVID-19. Conversely, of those who reported that the test showed they did have COVID-19, 22.9% (n=56) thought that they had not had COVID-19. Personal characteristics of participants are shown in Table 1.

**Table 1.** Table showing associations between participant personal characteristics and thinking you have had COVID-19.

| Participant characteristics | Level | Had COVID-19 |  | Odds ratio (95% CI) | Adjusted odds ratio (95% CI)<sup>†</sup> |
| --- | --- | --- | --- | --- | --- |
|  |  | Think have not had COVID-19 n=4656, n (%) | Think have had COVID-19 n=1493, n (%) |  |  |
| Gender | Male | 2197 (75.9) | 697 (24.1) | Reference | Reference |
|  | Female | 2459 (75.5) | 796 (24.5) | 1.02 (0.91 to 1.15) | 0.99 (0.87 to 1.12) |
| Age | 18 to 24 years | 1003 (70.5) | 419 (29.5) | Reference | Reference |
|  | 25 to 34 years | 823 (67.3) | 400 (32.7) | 1.16 (0.99 to 1.37) | 1.07 (0.89 to 1.28) |
|  | 35 to 44 years | 751 (71.9) | 294 (28.1) | 0.94 (0.79 to 1.12) | 0.80 (0.66 to 0.98) |
|  | 45 to 54 years | 554 (77.2) | 164 (22.8) | 0.71 (0.58 to 0.87)* | 0.62 (0.49 to 0.78)* |
|  | 55 years and over | 1525 (87.6) | 216 (12.4) | 0.34 (0.28 to 0.41)* | 0.36 (0.29 to 0.44)* |
| Have a child | No | 2005 (76.4) | 621 (23.6) | Reference | Reference |
|  | Yes | 2386 (75.5) | 776 (24.5) | 1.05 (0.93 to 1.19) | 1.30 (1.14 to 1.50)* |
| Employment status | Not working | 1714 (82.8) | 357 (17.2) | Reference | Reference |
|  | Working | 2871 (71.9) | 1124 (28.1) | 1.88 (1.65 to 2.15)* | 1.24 (1.05 to 1.46) |
| Working in key sector | No | 3105 (80.5) | 753 (19.5) | Reference | Reference |
|  | Yes | 1551 (67.7) | 740 (32.3) | 1.97 (1.75 to 2.21)* | 1.52 (1.32 to 1.75)* |
| Highest educational or professional qualification | GCSE/vocational/A-level/No formal qualifications | 3382 (76.1) | 1060 (23.9) | Reference | Reference |
|  | Degree or higher (Bachelors, Masters, PhD) | 1200 (74.3) | 415 (25.7) | 1.10 (0.97 to 1.26) | 1.10 (0.95 to 1.26) |
| Region | Midlands | 781 (75.7) | 251 (24.3) | Reference | Reference |
|  | South & East | 1369 (76.7) | 416 (23.3) | 0.95 (0.79 to 1.13) | 0.98 (0.81 to 1.19) |
|  | North | 1120 (77.0) | 335 (23.0) | 0.93 (0.77 to 1.12) | 0.95 (0.78 to 1.17) |
|  | London | 701 (70.1) | 299 (29.9) | 1.33 (1.09 to 1.62)* | 1.10 (0.89 to 1.36) |
|  | Wales, Scotland and Northern Ireland | 685 (78.1) | 192 (21.9) | 0.87 (0.70 to 1.08) | 0.89 (0.71 to 1.12) |

\*p≤.005
†Adjusting for all other personal characteristics

Younger participants, those who had a child, those who were employed (full-time, part-time, or self-employed), and those who worked in a key sector were more likely to report thinking that they had had COVID-19 (see Table 1).

### Differences between those who think they have and have not had COVID-19

18.5% of participants (n=1140) agreed or strongly agreed that they had some immunity to COVID-19. Those who thought they had had COVID-19 were more likely to agree that they had some immunity to COVID-19 (did not think they had had COVID-19: 10.7%, n=500; thought they had had COVID-19: 42.9%, n=640; see Table 2 and Figure 1).

**Table 2.** Table showing associations between thinking you have had COVID-19 and perceived immunity to COVID-19; worry about COVID-19; perceived risk of COVID-19 (to oneself and people in the UK); and total out-of-home activities in the last seven days (continuous outcomes).

| Participant characteristics | Level | Had COVID-19 |  | Unadjusted analyses |  |  | Adjusted analyses |  |  |
| --- | --- | --- | --- | --- | --- | --- | --- | --- | --- |
| | | Think have not had COVID-19<br>n=4656 | Think have had COVID-19<br>n=1493 | F | p | $\eta^2$ | F | p | $\eta^2$ |
| I think I have | 1=strongly | M=2.38, | M=3.33, | 998.11 | <.001* | .14 | 723.59 | <.001* | .11 |
| some immunity to COVID-19 | disagree to 5=strongly agree | SD=1.01 | SD=1.01 |  |  |  |  |  |  |
| Total out-of-home activity in the last seven days | Range=0 to 42 | M=6.69, SD=5.63 | M=9.35, SD=7.69 | 209.28 | <.001* | .03 | 83.70 | <.001* | .01 |
| Worry about COVID-19 | 1=not at all worried to 5=extremely worried | M=3.59, SD=1.01 | M=3.38, SD=1.12 | 50.16 | <.001* | .01 | 28.52 | <.001* | .01 |
| Perceived risk of COVID-19 to oneself | 1=no risk at all to 4=major risk | M=2.81, SD=0.76 | M=2.81, SD=0.76 | 0.01 | .93 | <.001 | 6.85 | .01 | .001 |
| Perceived risk of COVID-19 to people in the UK | 1=no risk at all to 4=major risk | M=3.39, SD=0.67 | M=3.30, SD=0.70 | 18.04 | <.001* | .003 | 5.67 | .02 | .001 |

\*p≤.005
†Adjusting for all personal characteristics and experimental condition

**Figure 1.**
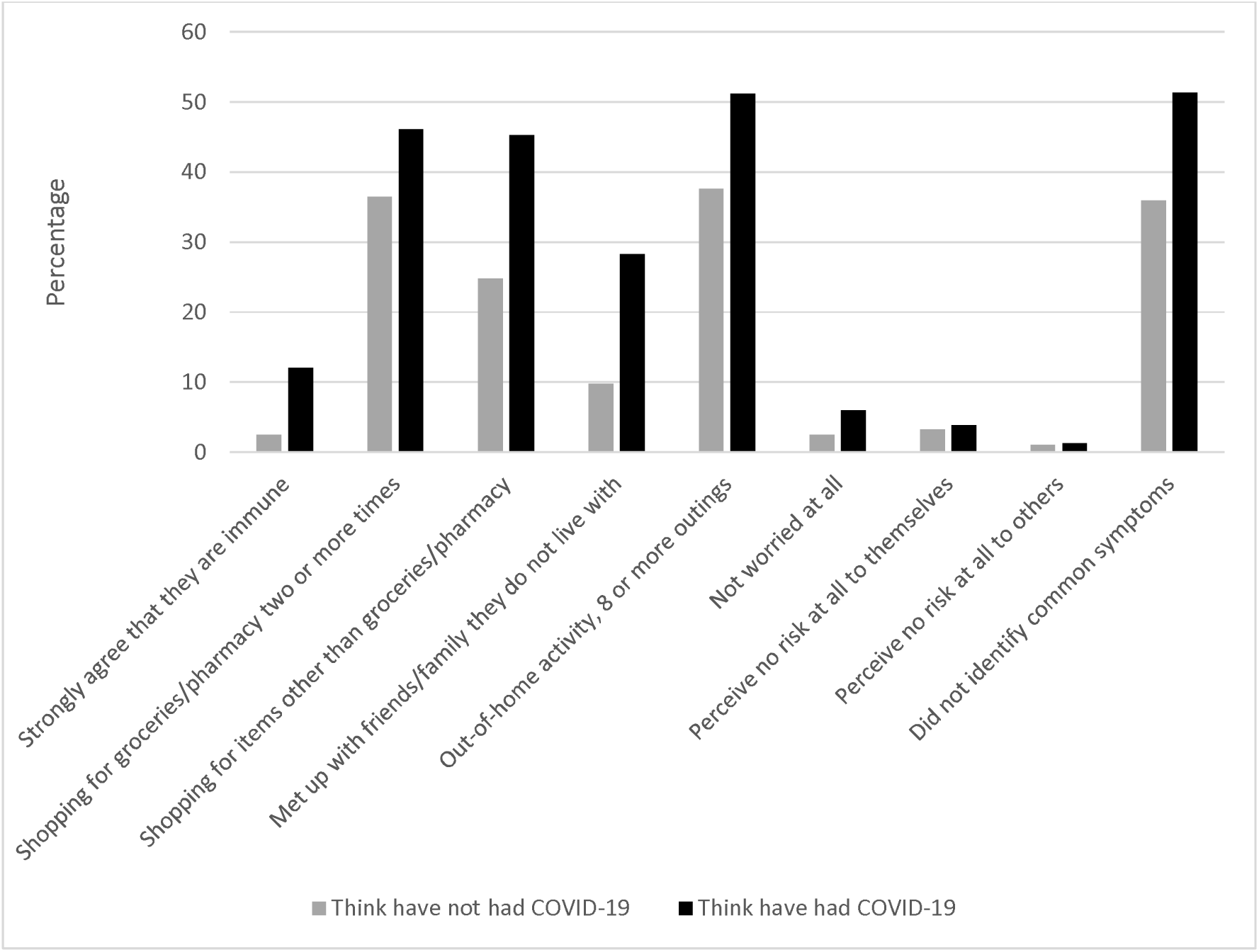
Graph depicting the percentage of people who: strongly agree that they are immune to COVID-19; went out to buy groceries/pharmacy on two or more days in the last seven days; went out to buy items other than groceries/pharmacy once or more in the last seven days; met up with friends and/or family they do not live with once or more in the last seven days; reported total out-of-home activity of eight or more (more than one outing per day on average); are not worried at all about COVID-19; perceive no risk at all to themselves from COVID-19; perceive no risk at all to people in the UK from COVID-19; did not identify cough and high temperature / fever as common symptoms of COVID-19 in those who think they have and have not had COVID-19.

In the last seven days, 38.9% (=2389) reported going out to the shops for groceries/pharmacy on two or more days; 29.8% (n=1833) reported going out to the shops for items other than groceries/pharmacy once or more; and 14.3% (n=878) reported meeting up with friends and/or family they did not live with once or more. Those who thought they had had COVID-19 were less likely to adhere to social distancing measures and went out shopping for groceries/pharmacy more frequently (Table 3). They also went out more times in total in the last seven days (Table 2).

**Table 3.**
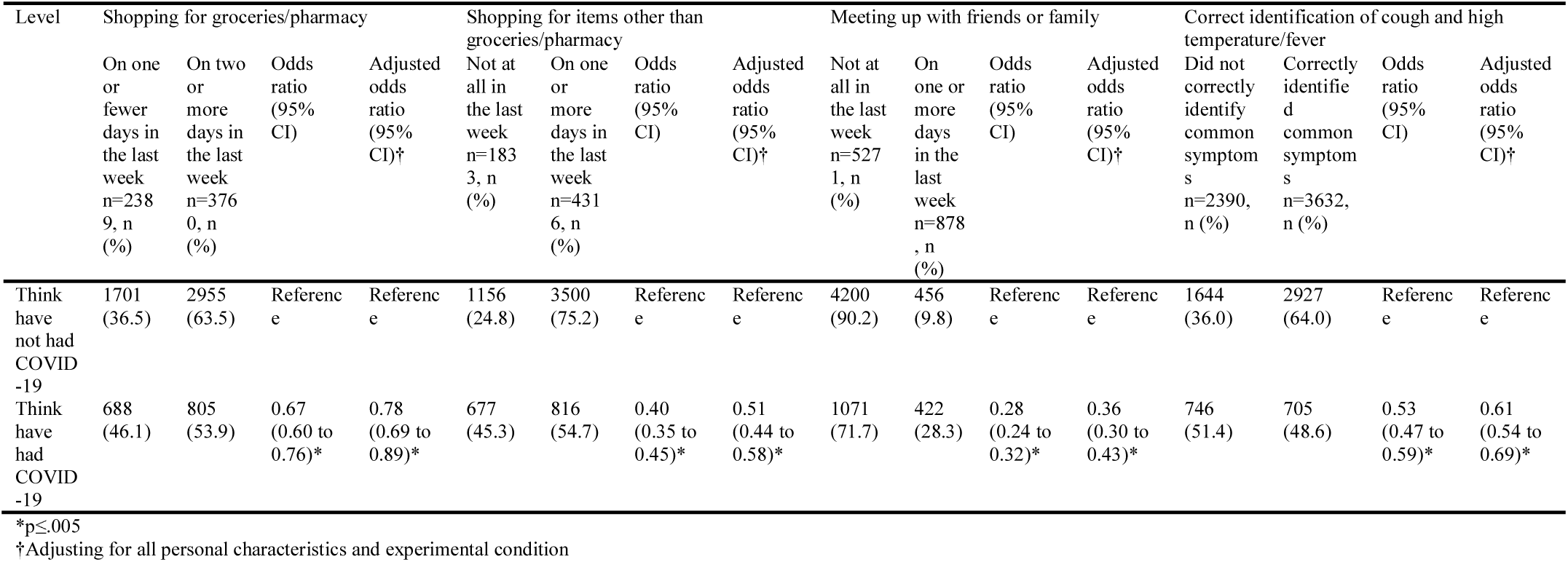
Table showing associations between thinking you have had COVID-19 and correct identification of most common symptoms of COVID-19; and adherence to social distancing measures (shopping for groceries/pharmacy, shopping for items other than groceries/pharmacy, and meeting up with friends and/or family who do not live with you; binary outcomes.

| Level | Shopping for groceries/pharmacy |  |  |  | Shopping for items other than groceries/pharmacy |  |  |  | Meeting up with friends or family |  |  |  | Correct identification of cough and high temperature/fever |  |  |  |
| --- | --- | --- | --- | --- | --- | --- | --- | --- | --- | --- | --- | --- | --- | --- | --- | --- |
|  | On one or fewer days in the last week<br>n=2389, n (%) | On two or more days in the last week<br>n=3760, n (%) | Odds ratio (95% CI) | Adjusted odds ratio (95% CI)† | Not at all in the last week<br>n=1833, n (%) | On one or more days in the last week<br>n=4316, n (%) | Odds ratio (95% CI) | Adjusted odds ratio (95% CI)† | Not at all in the last week<br>n=5271, n (%) | On one or more days in the last week<br>n=878, n (%) | Odds ratio (95% CI) | Adjusted odds ratio (95% CI)† | Did not correctly identify common symptoms<br>n=2390, n (%) | Correctly identified common symptoms<br>n=3632, n (%) | Odds ratio (95% CI) | Adjusted odds ratio (95% CI)† |
| Think have not had COVID-19 | 1701 (36.5) | 2955 (63.5) | Reference | Reference | 1156 (24.8) | 3500 (75.2) | Reference | Reference | 4200 (90.2) | 456 (9.8) | Reference | Reference | 1644 (36.0) | 2927 (64.0) | Reference | Reference |
| Think have had COVID-19 | 688 (46.1) | 805 (53.9) | 0.67 (0.60 to 0.76)* | 0.78 (0.69 to 0.89)* | 677 (45.3) | 816 (54.7) | 0.40 (0.35 to 0.45)* | 0.51 (0.44 to 0.58)* | 1071 (71.7) | 422 (28.3) | 0.28 (0.24 to 0.32)* | 0.36 (0.30 to 0.43)* | 746 (51.4) | 705 (48.6) | 0.53 (0.47 to 0.59)* | 0.61 (0.54 to 0.69)* |

\*p≤.005
†Adjusting for all personal characteristics and experimental condition

50.8% (n=3132) reported being very or extremely worried about COVID-19. Those who thought they had had COVID-19 were less worried about COVID-19 (see Table 2).

17.7% (n=1091) perceived a major risk of COVID-19 to themselves, while 47.0% (n=2893) perceived a major risk of COVID-19 to people in the UK. There was no evidence for an association between thinking you had had COVID-19 and perceived risk of COVID-19 (see Table 2).

59.1% (95% CI 57.8% to 60.3%, n=3632) correctly identified cough and high temperature / fever as two out of the three most common symptoms of COVID-19. Those who thought they had had COVID-19 were less likely to correctly identify these symptoms (see Table 3).

### Sensitivity analyses

Of those who had not been tested for COVID-19 (n=5574), 20.0% (95% CI 19.0% to 21.1%) thought they had had COVID-19 (n=1117).

In adjusted analyses, women were more likely to think that they had had COVID-19 (aOR=1.16, 95% CI 1.01 to 1.34). There was no evidence for an association between having a child or employment status and thinking you had had COVID-19.

There was also no evidence for an association between thinking you had had COVID-19 and: going shopping for groceries/pharmacy on two or more days in the last week, correct identification of two of the most common symptoms of COVID-19, and total out-of-home activity.

## DISCUSSION

Almost one quarter of participants thought they had had COVID-19. This percentage is higher to that seen in other surveys from the UK, with findings from daily tracker surveys indicating that approximately 10% to 18% think that they have had COVID-19.(8, 9) Differences in findings may be explained by the fact that these data only cover dates until 20^th^ April. Although we cannot be sure of the true proportion of the population that have had COVID-19, it is likely much lower, with an estimate by the Chief Medical Officer for England on 24^th^ April being “unlikely [to be] much above 10 per cent”.(10) It is likely that a substantial element of self-misdiagnosis underlies the high rate that we observed. This is supported not only by the high number of participants who felt they had had COVID-19 and who were unable to identify cough and fever as among the top three most common symptoms of the illness (52.8%) but also by the high proportion of people who reported that their test showed they did not have COVID-19 and who nonetheless believed they had had COVID-19. The proportion of the population who believe, rightly or wrongly, that they have had COVID-19 will only increase over time. Understanding how this affects behaviour is therefore important.

We found that people who thought they had had COVID-19 were more likely to think that they had some immunity to the virus and were less likely to adhere to social distancing measures. In particular, people were less likely to report adhering to measures that are not allowed at all in the UK, such as meeting up with friends and/or family that you do not live with and shopping for nonessentials. They also reported making more outings in the last week than those who did not think they had had COVID-19, however this result should be taken with caution as there was no longer an association in our sensitivity analyses. Given the cross-sectional nature of our data, it is impossible to be clear on causality – it may be that not adhering to social distancing rules leads to a greater likelihood of contracting COVID-19. However, the findings do fit with concerns expressed by the WHO that believing oneself to have had COVID-19 results in reduced adherence to protective behaviours.(3)

This finding has important implications at an individual level and also widely. Social norms are known to affect adherence to quarantine.(11) Therefore, people may be less likely to adhere to “lockdown” measures if they perceive other people, such as those who think they have had COVID-19, not to be adhering. To date, there are no communications specifically targeting those who think they have had COVID-19. This will become increasingly important in minimising transmission as the outbreak continues. Communications should acknowledge the growing proportion of the population who think that they have had COVID-19 and should issue targeted recommendations for this group explaining why it remains important to adhere to social distancing measures, maintain good hand hygiene and, for healthcare workers, use personal protective equipment where necessary.

In addition to associations with behaviour, thinking that you had had COVID-19 was associated with decreased worry about COVID-19. This appears logical, however there was little evidence for an association between perceived risk (to oneself and others) and believing you have had COVID-19. As we did not measure different factors that may contribute to worry (e.g. concern about personal finances / job, impact on physical / mental health), we are unable to tell which specific worries may be driving this decrease. It should be noted that differences detected in worry about COVID-19, and perceived risk of COVID-19 between those who did and did not think they had had the virus were small and may not be meaningful in real world situations.

Older participants were less likely to think they had had coronavirus. This may be because of a greater proportion of this group who are “shielding” (not leaving the home at all for at least 12 weeks). Those who have a child were more likely to report having had coronavirus perhaps linked to greater exposure, or perceived exposure, among this group.(12, 13) However, schools in the UK have been closed since 23^rd^ March 2020 except for children of key workers (14) reducing contacts between children(15) and these results pertaining to having children should be taken with caution as there was no longer any evidence for an association when analysing only those who had not been tested for COVID-19. Those who were employed (full-time, part-time or self-employed) were also more likely to think that they had had COVID-19, as were those working in key sectors. This may be due to increased objective exposure and perceived exposure as these groups continue to go out to work, while others work from home or stop working. There was no longer any evidence for an association between employment and thinking you had had COVID-19 when removing those who had been tested, therefore this interpretation should be taken with caution.

This study has several limitations. First, while quotas were used to ensure a sample that was broadly representative of the general UK population, we cannot be certain whether respondents in survey panels are representative of the general population.(16, 17) We also cannot rule out participation bias. Given potential participants were not aware of the topic of the survey before starting it, the risk of this is low. Second, we did not differentiate between outings that were in line with Government guidelines and those that were not in our measure of “total out-of-home activity”. Third, because we used a cross-sectional study design, we are unable to determine the direction of associations. Fourth, due to the large sample size, small differences between groups were statistically significant. Where detected differences were very small, there may not be meaningful influence of these differences (e.g. perceived risk to self).

People are likely to change their behaviour in line with their belief of whether they have had COVID-19. Even when tested, the reported result of an antigen test was not necessarily reflected in people’s belief about whether they had had COVID-19. Results from this study indicate that people who think they have had COVID-19 are less likely to adhere to social distancing measures. Clear, targeted communications might be used to advise this constantly growing group both to reduce reliance on self-diagnosis in the absence of a test and to provide advice on what behaviour changes, if any, are advisable.

## Data Availability

Anonymised data will be made available upon reasonable request.

## DATA SHARING STATEMENT

Anonymised data will be made available upon reasonable request.

## DISSEMINATION DECLARATION

Dissemination of study results to study participants is not possible due to the anonymous nature of data collection.

